# Climate Change and Eco-Anxiety in the US: Predictors, Correlates, and Potential Solutions

**DOI:** 10.1101/2022.08.28.22279314

**Authors:** Katherine Kricorian, Karin Turner

## Abstract

Climate change has many adverse human health effects, including increased anxiety. However, eco-anxiety may also motivate climate action. An online survey was developed and distributed to examine factors associated with eco-anxiety. Logistic regression analysis showed that significant predictors of eco-anxiety include greater media exposure to climate change information, more frequent discussions about climate change with friends and family, the perception that climate change will soon impact one personally, being younger, and being female. Additional analyses suggested that ecoanxiety was associated with a range of both positive and negative emotional impacts including motivation, interest, sadness, and tension. Eco-anxiety was also associated with greater likelihood to engage in environmental behaviors such as recycling. Volunteering for environmental causes and accessing straightforward information with less scientific jargon were found to have particular potential for anxiety reduction among the eco-anxious. The research suggests practical strategies to reduce eco-anxiety while retaining engagement in mitigating climate change.

## BACKGROUND

The World Health Organization calls climate change the biggest health threat that humanity faces today (World Health Organization, 2021). Beyond the immediate casualties caused by extreme weather events, climate change has also been associated with increased rates of allergies, asthma, heart attacks and strokes (Parker et al., 2019). Furthermore, Taylor (2020) discussed how climate change can increase both the likelihood of extreme weather events and pandemics, because warmer temperatures can create more hospitable habitats for disease vectors and viruses. People escaping extreme weather events may seek mass housing in shelters with other climate refugees, exacerbating the spread of contagious disease. Warmer temperatures and shorter winters allow insects carrying disease to breed for longer and travel further, facilitating insect-borne infectious disease outbreaks. For example, in Latin America alone, a 4°C increase in temperature is projected to increase the number of dengue fever cases by 8 million each year. Pollution has also been linked to various neurological and behavioral disorders, ranging from Alzheimer’s disease to autism spectrum disorders (Ramadan and Ataalah, 2021).

One often overlooked effect of climate change’s rising threat is that it can increase anxiety levels and worsen population mental health. Sociopolitical, economic and physical health problems caused by climate change, including forced migration, food shortages, job loss, increases in disease transmission, air pollution, and loss of social support can lead to mental illness (American Psychiatric Association, 2019). Exposure to natural disasters and severe weather events are correlated with anxiety, depression, sleep disorders, and post-traumatic stress disorder (PTSD) (Warsini et. al, 2014). Edwards, Gray and Borja (2021) found that experiencing natural disasters was associated with increased parental stress, food insecurity, and family violence, among other effects. Cruz et al. (2020) found specific extreme weather impacts associated with increased rates of psychological distress, anxiety, depression and PTSD, including whether the victims had experienced evacuation, displacement, disruption of essential services (e.g. electricity), and inability to attend work or school.

Ruminations about climate change’s future impacts can also damage mental health (Pihkala, 2018). “Eco-anxiety” is one term used to describe an emotional response to climate change caused by fear of the impact of climate change on one’s future (or the futures of loved ones), characterized by anxiety and stress. However, emotions related to climate change are nuanced and complex. Ramadan et al. (2021), identified ten different conceptualizations of the negative emotional response to climate change, including climate anxiety, climate change worry, eco-anxiety, and eco-paralysis. Stanley et al. (2021) further explored the differing implications and impacts of eco-depression, eco-anger, and eco-anxiety. Clayton and Karazsia (2020) developed a measure of climate change anxiety, including subscales for cognitive and functional impairments, and found correlations with general anxiety and depression. Eco-anxiety harms even people who have not personally experienced a climate crisis, causing trauma, stress, grief, depression, and general cognitive impairment (Dailianis, 2020). Hurley et al. (2022) described climate anxiety as a rational response given the enormous impacts of climate change, and Verplanken and Roy (2013) found that people who frequently worried about climate change were no more likely than infrequent ecological worriers to have pathological general levels of worry or emotional instability. In contrast, Reyes et al. (2021) found that higher climate change anxiety was associated with lower mental health in a sample of young adults from the Philippines.

Although eco-anxiety may be a reasonable and comprehensible reaction to climate change, it can still be distressing and damaging (Comtesse et al., 2021). Therapeutic strategies to address these issues have not been researched extensively, but include resilience training and climate awareness. One important public health challenge in treating eco-anxiety is helping people learn to cope with climate change without becoming apathetic, managing a society’s emotional responses at a complex level. Behavioral strategies may play a role in managing eco-anxiety while retaining some of its benefits, although these strategies have not been closely analyzed (Clayton, 2020). For example, some studies suggest the use of “social prescribing efforts” as solutions to climate anxiety. Social prescribing in general, such as the promotion of community-based activities including leisure, education and the arts, has been shown to have significant positive impact on mental and physical health (Munford et al., 2020). Social prescribing activities that may have a positive effect on eco-anxiety include spending time in nature and community-based work to support social connections and community-wide engagement, but these solutions have not yet been studied in-depth (Cunsolo et. al, 2020; Howarth et al., 2020).

Some studies suggest that eco-anxiety has not been associated with any particular behavioral responses to climate change (Clayton & Karazsia, 2020). However, some researchers suggest that stress about climate change motivates people and encourages climate action, hypothesizing that these effects may serve as potential positive aspects of eco-anxiety. Sanson and Bellemo (2021) stated that, when it comes to climate change, “action is the antidote to despair.” Verplanken and Roy (2013) found that habitual ecological worry was associated with positive environmental behaviors, such as recycling. Schwartz et al. (2022) determined that collective climate action may have a protective effect against climate anxiety, but did not find this result for individual environmental actions (such as personal recycling). In contrast, other studies suggest that eco-anxiety can engender feelings of “eco-paralysis,” or mental inhibition and levels of anxiety so extreme that they prevent one from taking action, resulting in immobilization or apathy (Albrecht, 2011; Usher, 2019).

One factor that may contribute to eco-anxiety is lack of information or overwhelming information about climate change and its effects. Exposure to climate change through media influences stress and mental health (American Psychiatric Association, 2019). Additionally, the indifference and ignorance of others to the climate crisis and lack of coordinated global action to mitigate it can cause great distress among those who care about the environment, leading to eco-anxiety (Rao and Powell, 2021). Frequent exposure to climate change information, such as through social media, newspapers, and television, has been correlated with heightened rates of eco-anxiety. Fritze et al. (2008) observed that climate change information can be so worrying that people react with skepticism and denial. Dodds (2021) noted that climate change doubt, denial and apathy serve as defense mechanisms against climate anxiety. Interestingly, research has also associated media exposure to climate change information with empowerment, indicating the potential additional motivational features of eco-anxiety. For example, Maran and Begotti (2021) found that climate anxiety in a sample of Italian students was positively correlated with both media exposure to climate change information and self-efficacy in dealing with climate change. Another study observed that newspapers and media sources in the United States are often selective in the content they portray, which can influence climate perceptions. Many newspapers portray teen activism against climate change in a negative light to appeal to adult readers, increasing eco-anxiety and sentiments that not enough work is being done or that the being done is ineffectual. Additionally, fear of being misrepresented in the media can repel climate activists from engaging productively with media, resulting in less coverage of climate change efforts (Duffy, 2022).

Eco-anxiety is also influenced by factors including socioeconomic status, geographic location, and culture, causing it to disproportionately impact different groups. Disadvantaged groups may be more likely to be exposed to environmental contaminants: Verbeek (2018) found that levels of air pollution were correlated with lower household incomes and higher unemployment levels. Those with a lack of access to resources and living in more vulnerable geographic conditions, such as in rural areas or closer to coasts, are more likely to experience climate anxiety (Cianconi et. al, 2020). Rural individuals, who may be more likely to labor in agriculture and thus be especially dependent on the environment, are more likely to experience climate-related insomnia, obsessive thinking, panic attacks, and appetite changes or eating disorders (Castellone, 2018). In addition to rural farmers, Indigenous Peoples, to whom the land has enhanced cultural significance, are also more likely to suffer from eco-anxiety (Cunsolo & Ellis, 2018; Middleton et. al, 2020).

Marginalized individuals such as people belonging to stigmatized racial, gender, and economic groups are more likely to suffer from the negative impacts of climate change, including mental illness (Hayes et. al, 2018). Cruz et al.’s (2020) analysis suggested that women, especially those in more vulnerable social positions, experienced greater negative mental health impacts of extreme weather events and that populations with unemployment or prior medical conditions are more likely to experience mental health declines after severe weather. Obradovich et al. (2018) found that climate change in the form of multiyear temperature increases and extreme weather events was associated with declines in mental health, with these effects greater for women and people with lower incomes.

Age also influences the mental health effects of climate change. Young people with developing brains, limited emotional resilience, and less experienced adapting to threats may be at greater risk for the effects of climate change and have more climate stress (Vergunst & Berry, 2021). Hickman et al. (2021) conducted a global survey of young people and found that more than half of 16-25 year olds were “very” or “extremely” worried about climate change and reported feeling sad, anxious, angry, powerless, helpless and guilty regarding climate change. Nearly half of young people surveyed said their feelings about climate change negatively affect their daily lives and three quarters were fearful of the future. Experiencing extreme stress about the climate can cause damage to brain chemistry, structure and function, creating the potential for even more serious mental illness later in life, including substance abuse and depression (Wu et. al, 2020). These age-related impacts of climate change may also have implications for the effects of climate anxiety on motivation and stress. Political orientation, eco-skepticism, and eco-anxiety may also be related. Cann, Weaver and Williams (2021) conducted social media sharing analysis of climate change information and found that levels of climate skepticism were associated with political ideology and information network polarization. As climate change continues to affect the planet, ecoanxiety and broad-scale research to understand the demographic and information exposure factors associated with it have increasing importance. It is critical that populations with high likelihood of suffering from eco-anxiety can be targeted for interventions, and additional exploration is needed regarding the efficacy of eco-anxiety interventions given that research in this area has been limited. Therefore, we conducted research to assess factors associated with climate change anxiety and potential solutions.

## METHODS

An English-language online survey questionnaire was developed and distributed to collect data about climate change attitudes. Institutional Review Board (IRB) exemption was received from WCG IRB, an IRB specializing in medical and public health research reviews. The IRB classified the survey as exempt because it was an anonymous survey and no personally-identifying information (PII) was collected from respondents. The survey instrument was developed based on review of the literature and previously published surveys, and programmed using AYTM online survey development and deployment software. AYTM is a research software company that maintains a survey panel of over 100 million potential respondents in 40 countries.

Potential US respondents were invited by text message and email to participate in an online, English-language survey to collect data about climate change attitudes. These potential respondents had previously agreed to participate in surveys for research purposes. In January 2022, approximately N=5,000 potential respondents were invited to participate in the survey and a sample of N=2,000 US adults aged 18+ completed the survey, resulting in a response rate of 40%. At the beginning of the survey, respondents were informed of the purposes of the study and agreed to participate voluntarily. Formal consent was not obtained because survey data was collected anonymously. Survey respondents consisted of a broad range of ages, races, ethnic backgrounds, income levels and geographic origins across the US.

The survey aimed to assess factors associated with eco-anxiety and potential interventions to reduce its negative effects on mental health and quality of life while retaining engagement and motivation regarding climate change. At the beginning of the survey, respondents were asked to indicate their age and their country of residence and to provide their consent to participate in a survey about climate change. People who were under age 18 and/or currently lived outside the US were thanked and terminated from the study.

In the first portion of the survey, respondents were asked to indicate their knowledge, beliefs, behavior and emotions regarding climate change. The complexity and measurement of the negative emotional responses to climate change has been the subject of much exploration and discussion. Measures used were informed by several scholarly approaches, including those of Clayton and Karazsia (2020), Coffey et al., (2021), Helm et al., (2018), Hogg et al., (2021), Stanley et al. (2021) and Stewart (2021). In the next section of the survey, respondents answered questions regarding their demographics. At the end of the survey, respondents were thanked for their participation. Data were analyzed using SPSS v.28.0 statistical analysis software. First, logistic regression analysis was done to understand predictors of climate anxiety. Then, *χ*^2^ tests were conducted to examine the relationship of climate anxiety with attitudes and behavior. In both the logistic regression and the χ^2^ tests, analysis focused on self-reported responses to the question “Does climate change make you feel anxious?”

## RESULTS

The survey sample consisted of people with a range of ages, relationship statuses, education levels and racial/ethnic backgrounds (see Table 1). More females than males completed the survey, but other demographic characteristics such as race and ethnicity mirrored US population values according to the US Census. More than one quarter of survey respondents (26%) indicated they were anxious about climate change, and the remainder did not (74%) (see Figure 1).

**Table 1:** Survey sample characteristics.

| Gender | Percent | Education Level | Percent | Relationship Status | Percent |
| --- | --- | --- | --- | --- | --- |
| Female | 57.1% | No college | 17.0% | Married | 41.5% |
| Male | 43.0% | Some college | 26.7% | Single | 34.1% |
|  |  | 2yr degree | 12.0% | Divorced | 10.5% |
|  |  | 4yr degree | 27.8% | Living with a significant other | 7.2% |
|  |  | Grad school degree | 10.8% | Widowed | 3.9% |
|  |  | Professional degree | 5.9% | Engaged | 1.4% |
|  |  |  |  | It's complicated | 1.6% |
| Race/ Ethnicity |  | Age |  | Annual Household Income |  |
| White / Caucasian | 63.4% | 18-24 | 7.2% | \$0 - \$25,000 | 26.4% |
| Black / African-American | 14.1% | 25-34 | 16.3% | \$25,000 - \$50,000 | 26.6% |
| Hispanic / Latino / Spanish | 12.4% | 35-44 | 20.2% | \$50,000 - \$75,000 | 20.0% |
| Asian | 5.7% | 45-54 | 18.8% | \$75,000 - \$100,000 | 13.0% |
| Multiracial | 2.0% | 55-64 | 20.2% | \$100,000 - \$200,000 | 12.2% |
| Native American | 0.7% | 65-74 | 15.2% | \$200,000 - \$500,000 | 1.9% |
| Other Race | 1.9% | 75+ | 2.3% | >\$500,000 | 0.2% |
| Parental Status |  | Mean Age | 47.9 years (SD=15.4) |  |  |
| No children | 71.7% | Median Age | 48.0 years |  |  |
| 1 child | 13.0% | Age Range | 18-90 years |  |  |
| 2 children | 9.5% |  |  |  |  |
| 3 children | 4.3% |  |  |  |  |
| 4 children | 1.1% |  |  |  |  |
| 5+ children | 0.6% |  |  |  |  |

**Figure 1:**
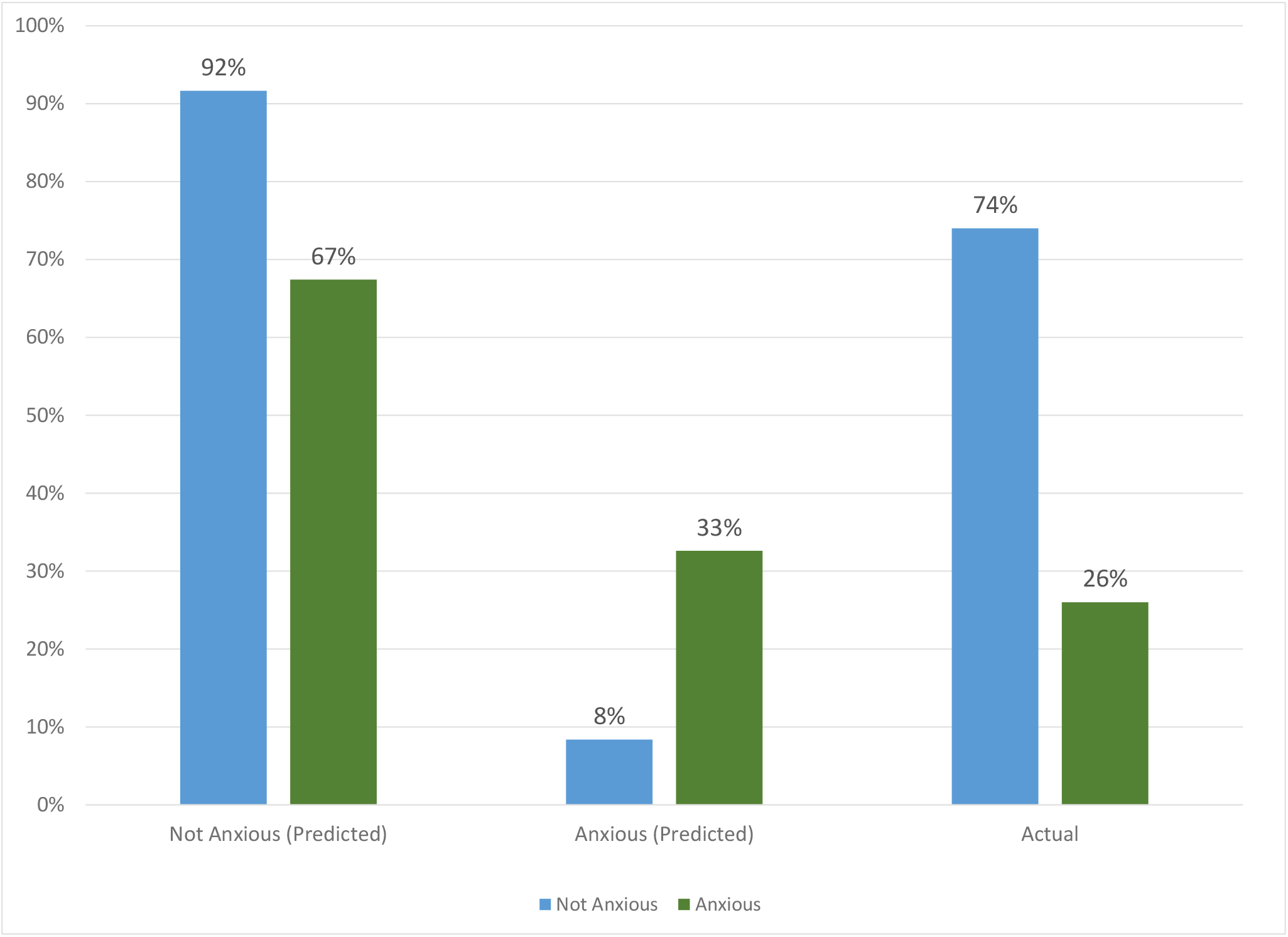
Logistic regression classification accuracy.

### ECO-ANXIETY PREDICTORS

In the logistic regression model, climate anxiety served as the dependent variable and demographics and climate change information exposure were independent variables. The model correctly classified 92% of those without eco-anxiety, substantially higher than the 74% correct classification expected by the null model. The model also correctly classified 33% of those with eco-anxiety, significantly better than the 26% correct classification expected by the null model (see Figure 1). The overall model had a significant fit at p<.0001. The Nagelkerke R_2_=.296, so about one third of the variation in the data was explained by the model (see Table 2). This is a moderately strong result for a social science model.

**Table 2:** Logistic regression results and model fit.

| | B | S.E. | Wald $\chi^2$ | df | Sig. | Odds ratio |
| --- | --- | --- | --- | --- | --- | --- |
| **How familiar are you with climate change?** |  |  | 1.621 | 4 | 0.805 |  |
| Very | 0.006 | 0.232 | 0.001 | 1 | 0.980 | 1.006 |
| Somewhat | 0.133 | 0.250 | 0.281 | 1 | 0.596 | 1.142 |
| Not very | 0.205 | 0.458 | 0.199 | 1 | 0.655 | 1.227 |
| Not at all | 1.089 | 0.957 | 1.295 | 1 | 0.255 | 2.971 |
| **How often do you hear about climate change in the media?** |  |  | 22.665 | 8 | 0.004 |  |
| 2-3 times per week | -0.028 | 0.262 | 0.012 | 1 | 0.914 | 0.972 |
| About once a week | -0.448 | 0.289 | 2.395 | 1 | 0.122 | 0.639 |
| 2-3 times per month | -0.962 | 0.339 | 8.044 | 1 | 0.005 | 0.382 |
| About once a month | -0.097 | 0.364 | 0.071 | 1 | 0.790 | 0.908 |
| Every 2-3 months | -1.894 | 0.677 | 7.837 | 1 | 0.005 | 0.150 |
| Several times a year | -1.054 | 0.513 | 4.220 | 1 | 0.040 | 0.348 |
| Once a year or less | -0.120 | 0.715 | 0.028 | 1 | 0.867 | 0.887 |
| Never | -1.909 | 1.094 | 3.043 | 1 | 0.081 | 0.148 |
| **How often do you discuss climate change with friends, family or other people you know?** |  |  | 26.866 | 8 | 0.001 |  |
| 2-3 times per week | 0.967 | 0.475 | 4.145 | 1 | 0.042 | 2.629 |
| About once a week | 0.398 | 0.474 | 0.706 | 1 | 0.401 | 1.490 |
| 2-3 times per month | 0.709 | 0.478 | 2.207 | 1 | 0.137 | 2.033 |
| About once a month | -0.103 | 0.496 | 0.044 | 1 | 0.835 | 0.902 |
| Every 2-3 months | 0.531 | 0.530 | 1.004 | 1 | 0.316 | 1.701 |
| Several times a year | 0.422 | 0.524 | 0.649 | 1 | 0.421 | 1.526 |
| Once a year or less | -0.091 | 0.549 | 0.028 | 1 | 0.868 | 0.913 |
| Never | -0.628 | 0.521 | 1.453 | 1 | 0.228 | 0.534 |
| **Generally speaking, do you usually think of yourself as:** |  |  | 3.551 | 3 | 0.314 |  |
| Republican | -0.445 | 0.242 | 3.374 | 1 | 0.066 | 0.641 |
| Independent | -0.115 | 0.198 | 0.336 | 1 | 0.562 | 0.891 |
| Something else (Not Democrat, Republican or Independent) | -0.245 | 0.296 | 0.684 | 1 | 0.408 | 0.783 |
| **When do you think climate change will start harming you and your friends/family?** |  |  | 47.850 | 8 | 0.000 |  |
| In 1-5 years | -0.437 | 0.233 | 3.517 | 1 | 0.061 | 0.646 |
| In 6-10 years | -0.948 | 0.352 | 7.234 | 1 | 0.007 | 0.388 |
| In 11-15 years | -1.011 | 0.513 | 3.885 | 1 | 0.049 | 0.364 |
| In 16-25 years | -1.817 | 0.642 | 7.994 | 1 | 0.005 | 0.163 |
| In 26-50 years | -0.913 | 0.469 | 3.795 | 1 | 0.051 | 0.401 |
| In 51-100 years | -0.854 | 0.614 | 1.934 | 1 | 0.164 | 0.426 |
| More than 100 years from now | -1.203 | 0.476 | 6.380 | 1 | 0.012 | 0.300 |
| Never | -3.143 | 0.611 | 26.436 | 1 | 0.000 | 0.043 |
| **Age** | -0.011 | 0.005 | 4.421 | 1 | 0.036 | 0.989 |
| **Gender(Male)** | -0.385 | 0.169 | 5.173 | 1 | 0.023 | 0.680 |
| **Highest level of education completed** |  |  | 6.168 | 5 | 0.290 |  |
| Some college | -0.048 | 0.272 | 0.031 | 1 | 0.859 | 0.953 |
| 2yr degree | -0.171 | 0.317 | 0.292 | 1 | 0.589 | 0.843 |
| 4yr degree | 0.267 | 0.277 | 0.926 | 1 | 0.336 | 1.305 |
| Grad school degree | -0.370 | 0.351 | 1.114 | 1 | 0.291 | 0.691 |
| Professional degree | -0.253 | 0.410 | 0.382 | 1 | 0.536 | 0.776 |
| **Constant** | 0.420 | 0.543 | 0.599 | 1 | 0.439 | 1.522 |

**Omnibus Tests of Model Coefficients**
|  | square | df | Sig. |
| --- | --- | --- | --- |
| Step | 228.213 | 38 | 0.000 |
| Block | 228.213 | 38 | 0.000 |
| Model | 228.213 | 38 | 0.000 |

**Model Summary**
|  | -2 Log likelihood | Snell R Square | Nagelkerke R Square |
| --- | --- | --- | --- |
|  | 938.305<sup>a</sup> | 0.204 | 0.296 |

Factors including familiarity with climate change, political party, and education did not serve as significant predictors of climate anxiety in the logistic regression model (see Table 2). Frequency of media exposure about climate change was associated with climate anxiety: in general, less frequent self-reported media exposure was associated with lack of climate anxiety. Similarly, infrequency of climate discussions with friends and family was associated with lack of climate anxiety. People who felt it would be a long time before climate change impacted them personally were less likely to be anxious, as were males and older people.

### ECO-ANXIETY CORRELATES

Associations between eco-anxiety and respondents’ attitudes and behavior was examined using *χ*^2^ analysis. Those who indicated they were not anxious about climate change were significantly more likely than those with eco-anxiety to say that climate change is harmless, a hoax, not going to affect my daily life, and exaggerated (all comparisons p<.05, see Figure 2). People without eco-anxiety were also significantly more likely to say that climate change is caused mostly by natural changes in the environment (28.2% vs. 15.8%, p<.05). Those with eco-anxiety were significantly more likely than those without eco-anxiety to say climate change is something almost all scientists agree about, is caused mostly by human activities, is real, and is already happening (all comparisons p<.05). Interestingly, only about half (52.3%) of those without climate anxiety said they believe climate change is real.

**Figure 2:**
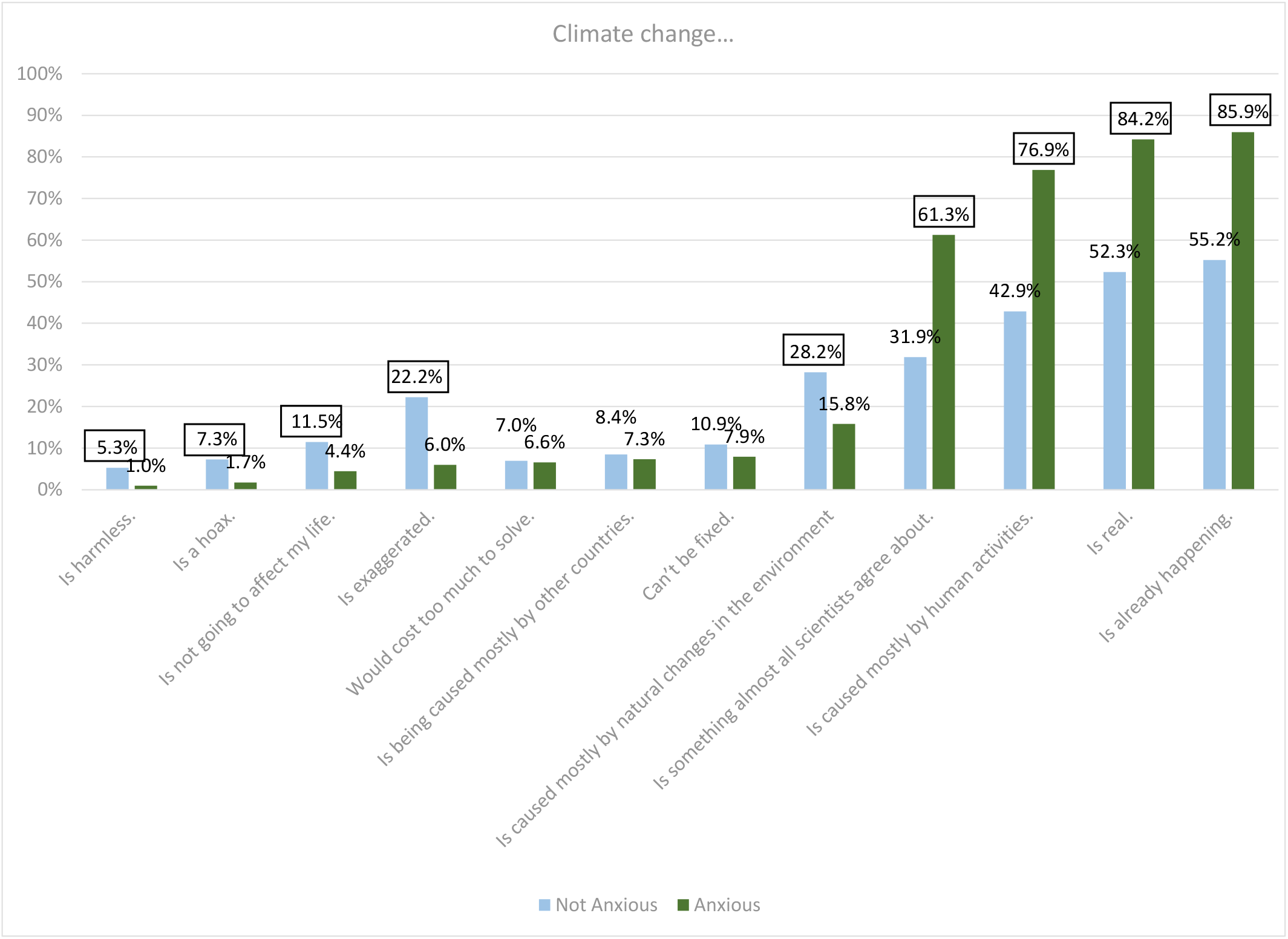
Beliefs about climate change. Survey question:*Select all of the statements you agree with. Climate change ….* Boxed numbers on graph are higher than comparison group at p<.05.

People without eco-anxiety were significantly more likely to say that information about climate change is fake, one-sided, not trustworthy, and too political (all comparisons p<.05, see Figure 3). More than a third (35.2%) of those without ecoanxiety said information about climate chance is too political vs. 25.0% of those with eco-anxiety (p<.05). Those with climate change anxiety were significantly more likely to say that information about climate change is hard to understand, confusing and overwhelming (p<.05). Nearly half (47.0%) of climate anxious respondents said climate change information is overwhelming vs. just 19.9% of respondents without climate anxiety (p<.05).

**Figure 3:**
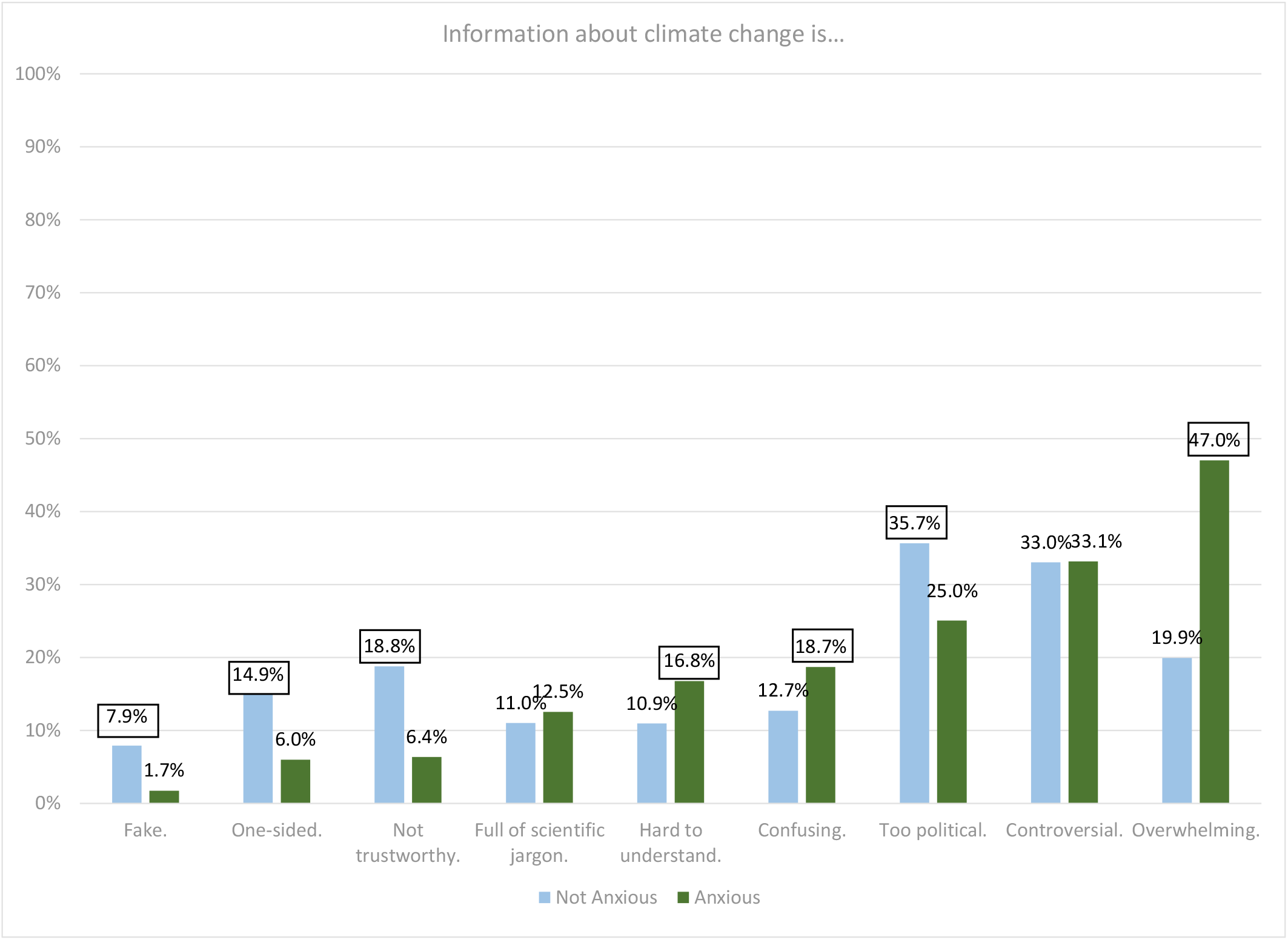
Perceptions of information about climate change. Survey question: *Please select all the statements you agree with. Information about climate change is … Boxed numbers on graph are higher than comparison group at p<.05.*

Respondents were also asked to indicate whether they felt other emotions about climate change. People without eco-anxiety were more likely to say they felt skeptical (16.2% vs. 8.9%, p <.05, see Figure 4). People with eco-anxiety were more likely to report a range of emotions: they were more likely to report being motivated and interested regarding climate change, but they were also more likely to say they felt depressed, angry, sad, overwhelmed, frustrated and worried (all comparisons p<.05). The survey also asked respondents to report how climate change is affecting their mental health, if at all. The results showed that 84.7% of those who were not anxious about climate change said their mental health was unaffected by climate change vs. 48.4% of those with climate change anxiety (p<.05). Nearly half (45.5%) of those with climate change anxiety said climate change made their mental health somewhat worse (vs. only 11.5% of those without climate change anxiety, p<.05) and 6.2% (vs. 3.8%, p<.05) said climate change made their mental health a lot worse (see Figure 5). People with climate change anxiety were significantly more likely to indicate that their ecoanxiety negatively impacted their wellbeing and quality of life. They were more likely to say that their concerns about climate change made it harder to get school/work assignments done (5.2% vs. 2.8%), have fun with family and friends (7.5% vs. 2.6%), and concentrate (10.2% vs. 4.2%) (all comparisons p<.05) (see Figure 6).

**Figure 4:**
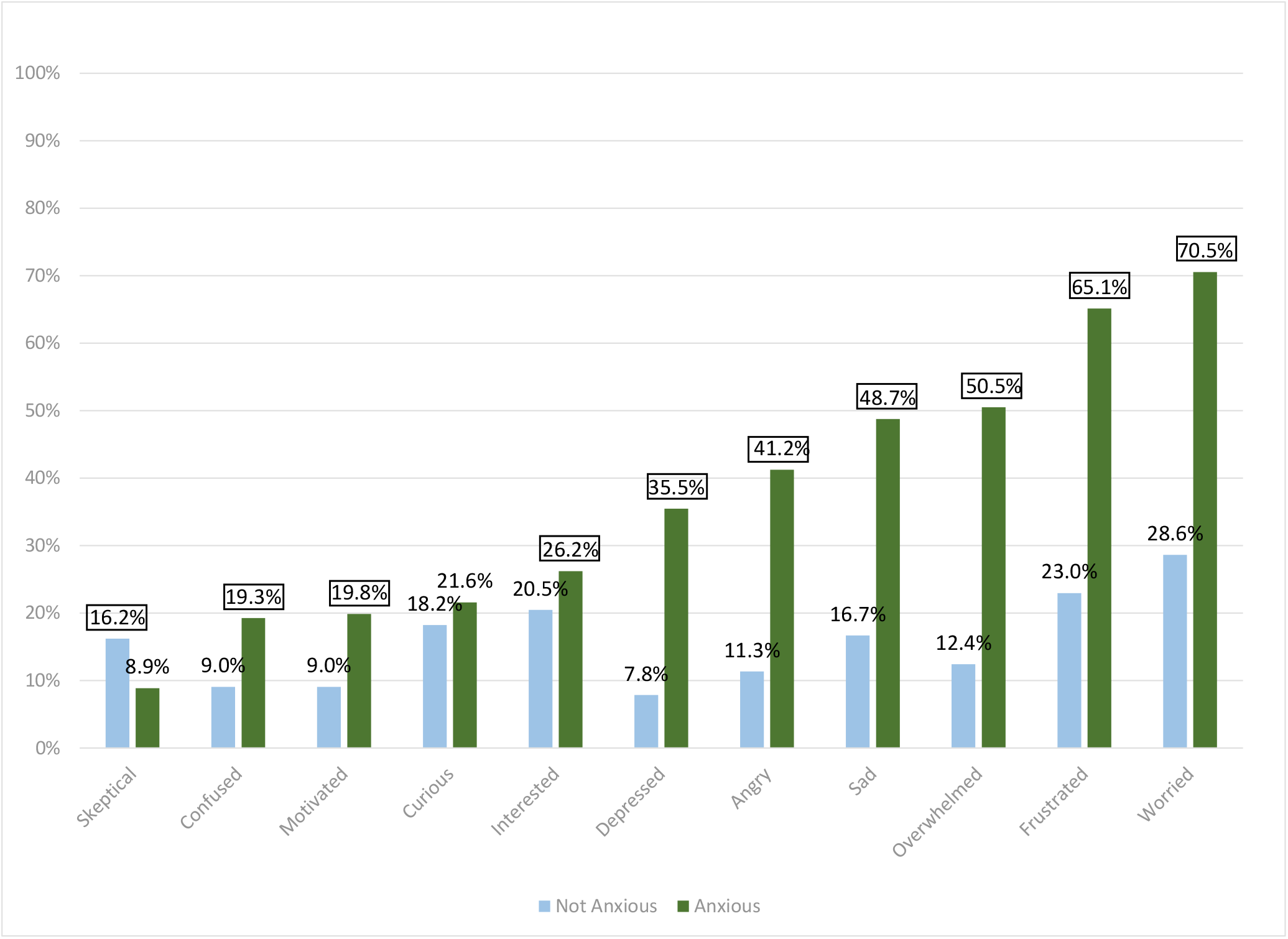
Feelings about climate change. Survey question: *Which, if any, of the following words describe how climate change makes you feel? Select all that apply.* Boxed numbers on graph are higher than comparison group at p<.05.

**Figure 5:**
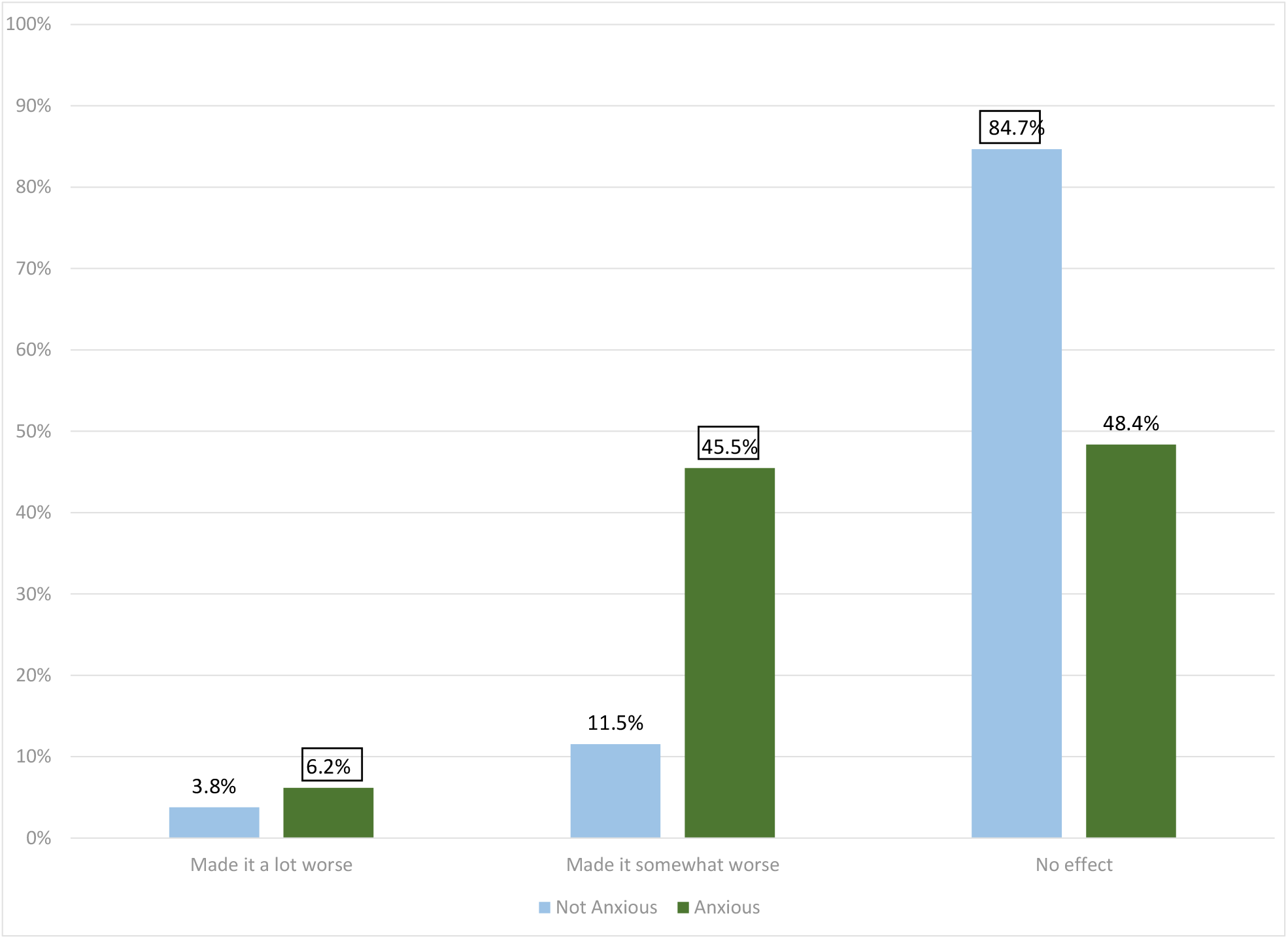
Climate change impact on mental health. Survey question: *How has climate change affected your mental health, if at all?* Boxed numbers on graph are higher than comparison group at p<.05.

**Figure 6:**
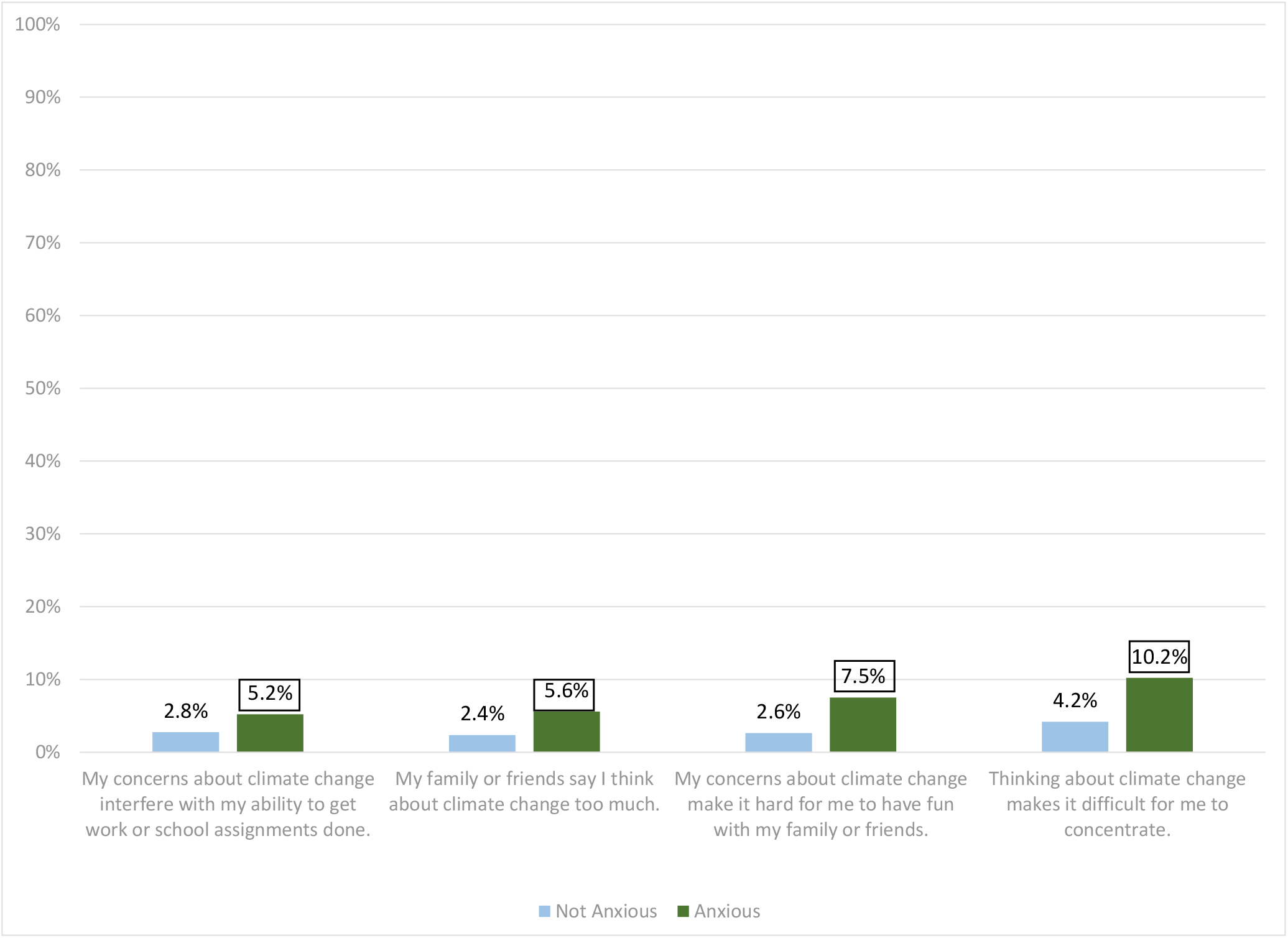
Cognitive and functional impairments due to eco-anxiety. Survey question: *Select all of the statements you agree with regarding climate change.* Boxed numbers on graph are higher than comparison group at p<.05.

### POTENTIAL SOLUTIONS

People with climate change anxiety were significantly more likely to exhibit environmentally conscious behaviors, such as being vegetarians, driving an electric/hybrid car, using public transportation, voting with relation to climate issues, recycling, reducing energy usage, and having confidence that their actions can have an impact on climate change (p<.05, see Figure 7). People with climate anxiety were more likely to support organizations and charities related to the environment or the effects of climate change through donating time or money (24.7% vs. 10.2%, p<.05, see Figure 8). The eco-anxious were also significantly more likely to support many other types of charitable organizations, such as animal charities, human and social services and health charities (p<.05). However, people without climate change anxiety were significantly more likely to support religious organizations (19.3% vs. 14.6%, p<.05). Of respondents not involved with environmental organizations, those with climate anxiety were more likely to cite structural barriers like lack of time, access, awareness, or experience (comparisons p<.05, see Figure 9). People without eco-anxiety were more likely to say that lack of interest was the reason they do not volunteer for or donate to environmental causes (17.0% vs. 6.4%, p<.05).

**Figure 7:**
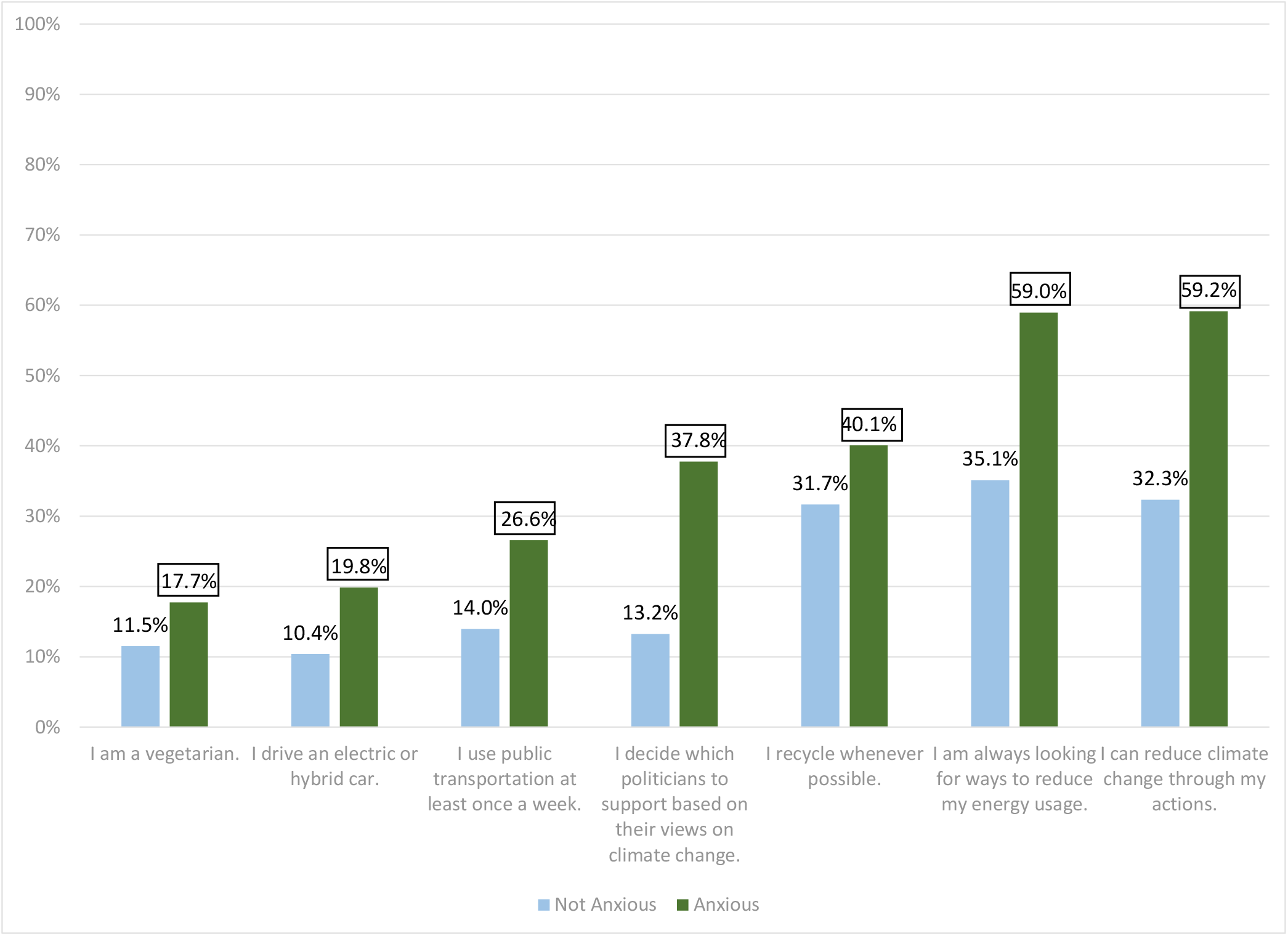
Environmental behaviors. Survey question: *Please select the statements that describe you. Select all that apply.* Boxed numbers on graph are higher than comparison group at p<.05.

**Figure 8:**
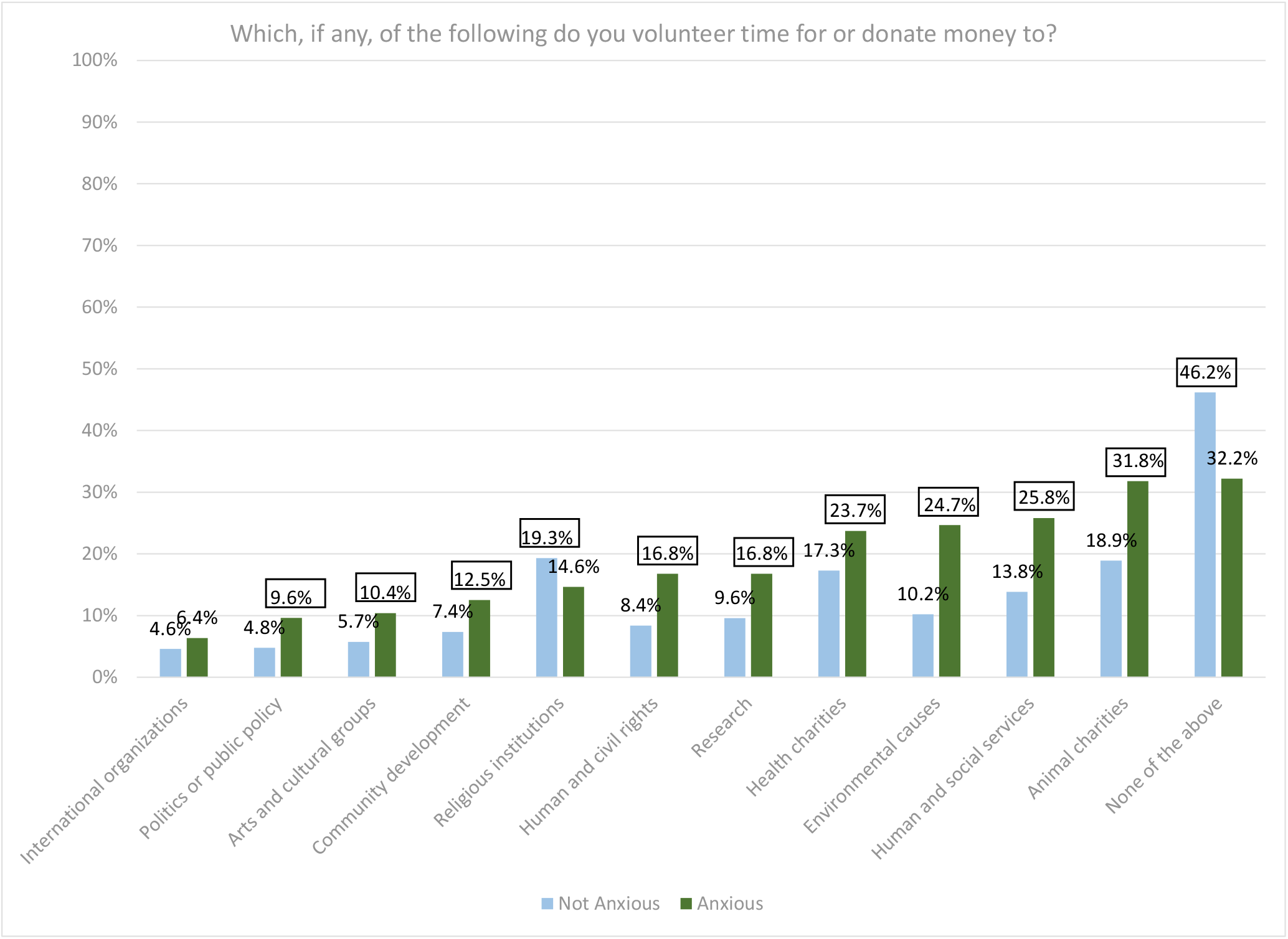
Current charitable behavior. Survey question: *Which, if any, of the following do you volunteer time for or donate money to? Select allthat apply.* Boxed numbers on graph are higher than comparison group at p<.05.

**Figure 9:**
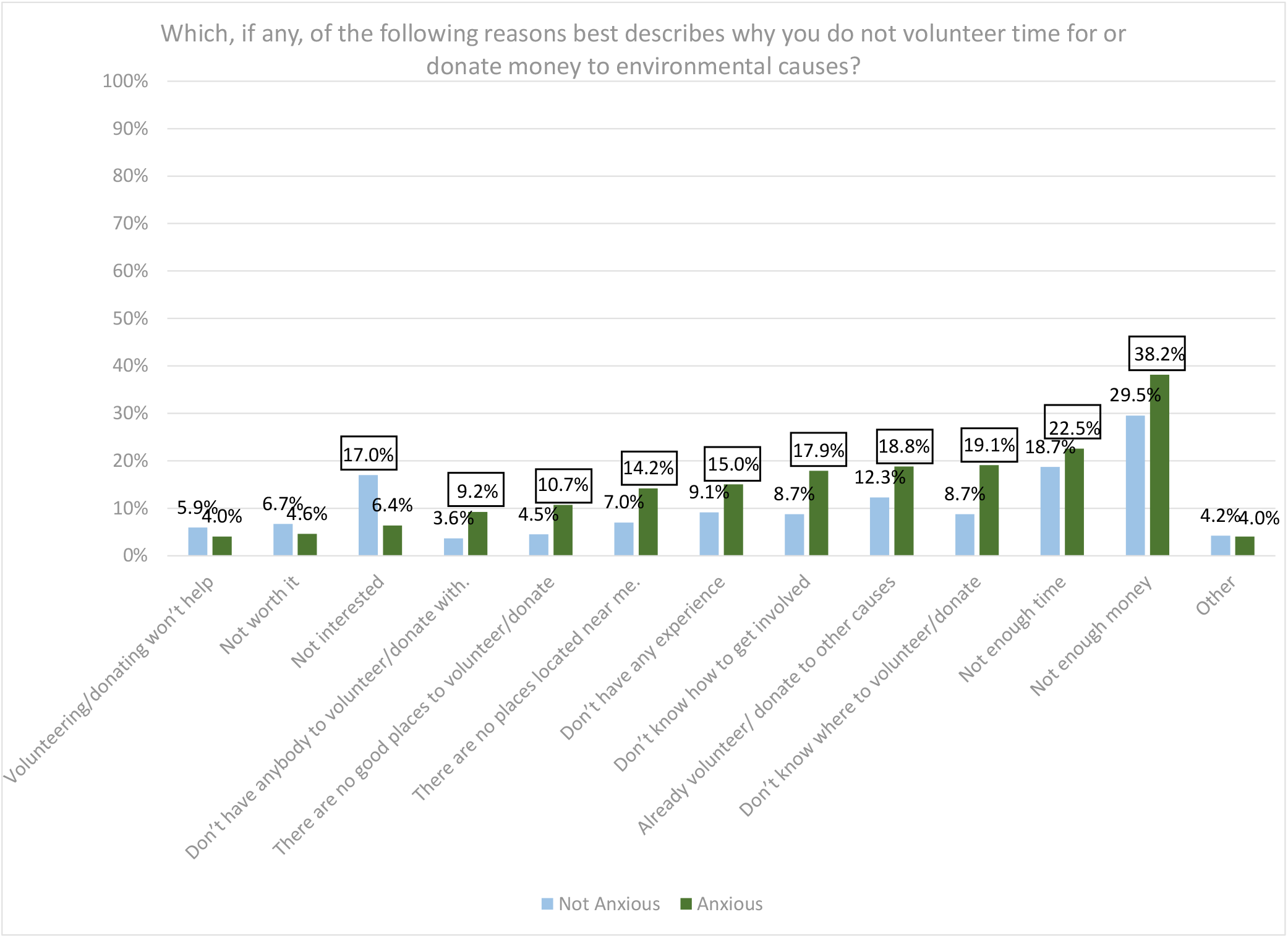
Reasons respondent does not volunteer for or donate to environmental organizations. Survey question (Asked only of those who indicated they did not volunteer for or donate to environmental causes): *Which, if any, of the following reasons best describes why you do not volunteer time for or donate money to environmental causes?* Boxed numbers on graph are higher than comparison group at p<.05.

Those without climate anxiety were more likely to say that climate anxiety would be reduced by less emphasis on climate change among government leaders and less media coverage of climate change (both comparisons p<.05, see Figure 10). People with climate anxiety were more likely to state that their stress would be reduced by personal actions such as donating money or time to environmental causes, learning more about current climate change reduction efforts, and learning how they can help climate change efforts (all comparisons p<.05) Those with climate anxiety were also more likely to cite the importance to institutional efforts in reducing eco-anxiety, including the need for additional laws to mitigate climate change and more scientific research into renewable energy sources (comparisons p<.05).

**Figure 10:**
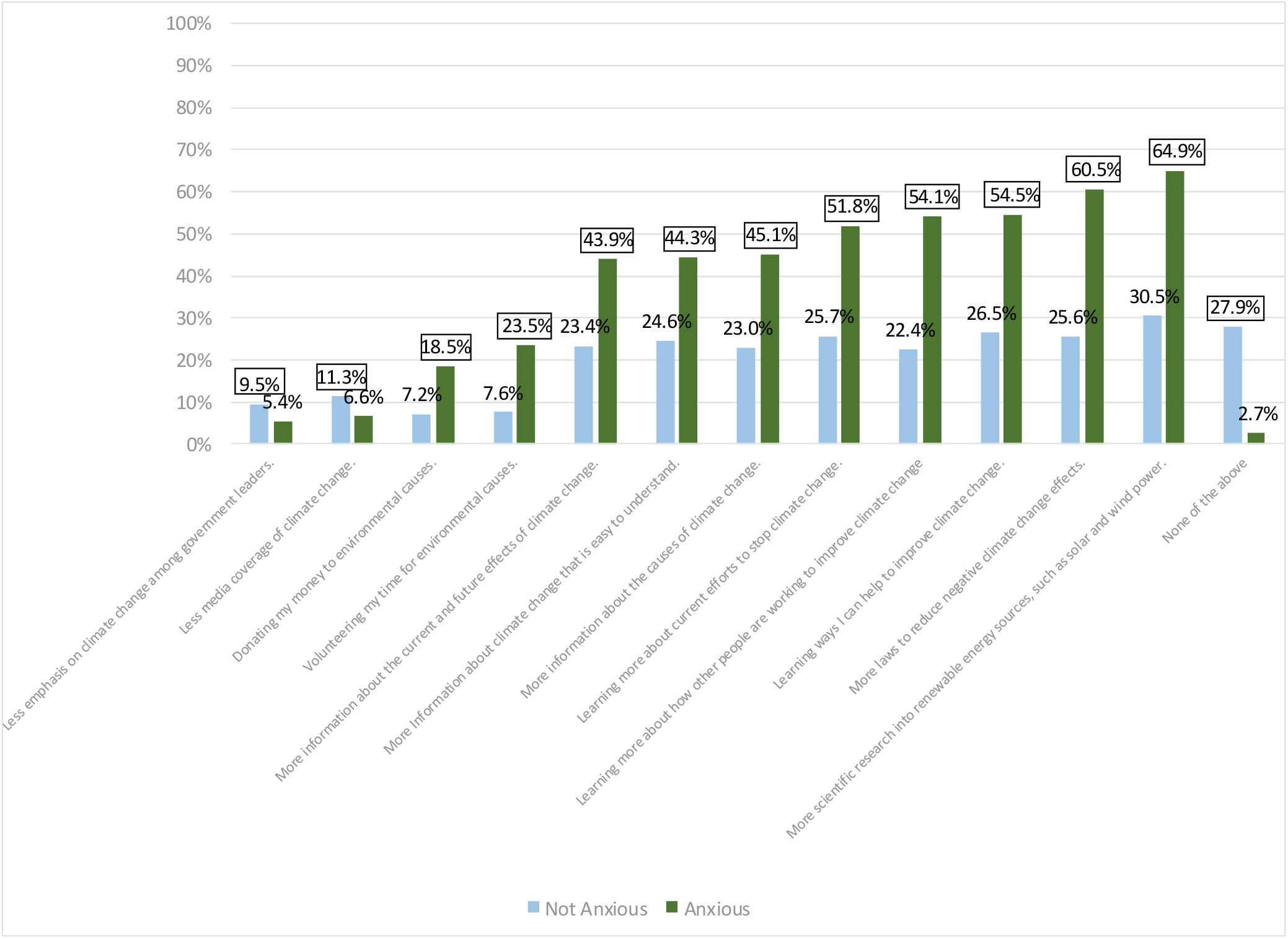
Factors that would make respondent feel less worried or anxious about climate change. Survey question: *Which, if any, of the following would make you feel less worried or anxious about climate change? Select all that apply.* Boxed numbers on graph are higher than comparison group at p<.05.

## DISCUSSION

Based on the logistic regression model results, climate change anxiety was associated with greater self-reported frequency of hearing about climate change in the media and discussing climate change with friends and family. However, the direction of causality was not clear: media exposure to climate change information and talking about it might cause climate change anxiety, or people who are already worried about climate change might seek out information and want to talk about it with others. Younger age and female gender were also associated with greater likelihood of climate change anxiety. Furthermore, people who thought climate change would happen further into the future were less likely to have climate change anxiety. These results support prior research that has shown that females have higher baseline levels of general anxiety, which may contribute to their likelihood to develop climate change anxiety (Bahrami & Yousefi, 2011). Young people, especially if they expect climate change to cause damage in the shorter term future, would logically be more anxious about climate change because they may anticipate that their lives overall will be more affected by it than older people, since young people expect to have more remaining years of life. This indicates that climate anxiety has several demographic factors that must be recognized when developing tailored, targeted solution strategies.

Some variables in the logistic regression did not have the expected relationship with eco-anxiety. Self-defined familiarity with climate change was not significantly associated with climate change anxiety. It was hypothesized that the greater knowledge that people felt they had about climate change, the more anxious they would be. The model results did not support this hypothesis. Political party affiliation and education levels also did not have significant relationships with climate change anxiety. Prior research would suggest that more politically conservative and less educated survey respondents would be less likely to have climate change anxiety, but these results were not found in the analysis.

The results identified both positive and negative correlates of climate change anxiety. People with eco-anxiety were more likely to say that climate change makes them feel hopeless, tense, and troubled and made their mental health either a lot or somewhat worse. Additionally, people with eco-anxiety were more likely to report that their climate concerns damaged their ability to work productively, concentrate, and enjoy free time with friends and family. This indicates the damaging toll that eco-anxiety may take on personal wellbeing. However, the research also revealed several potentials to utilize eco-anxiety and produce a positive impact. For example, those with climate change anxiety were also more likely to say they take action in their daily lives to reduce their contributions to climate change, being more likely to look for ways to reduce energy usage, recycle, and use public transportation. Those with eco-anxiety were more likely to say that climate change makes them feel motivated and interested and were more likely to volunteer and/or donate to environmental organizations. Because ecoanxiety is so complex, with both positive and negative aspects, it is important to develop and identify solution strategies that will exert the greatest impact on bettering mental health while fighting apathy and retaining engagement in the climate cause.

To this end, the research identified several solutions to climate change anxiety, including behavioral strategies, a potential indicated by previous studies. Those with anxiety were more likely to believe that environmental volunteering would reduce their anxiety. This may be because volunteering offers opportunities to directly impact the local environment and help volunteers have a personal impact on the cause, which may soothe anxiety about lack of involvement or feelings of helplessness. Those who did not volunteer cited reasons such as a lack of experience or awareness of how to get involved. To combat this, environmental organizations may have an opportunity to promote volunteering opportunities and organize easy-to-access climate volunteering events for various communities. Those with eco-anxiety were also more likely to say more information on climate change causes, effects, and current mitigation efforts would reduce their anxiety, as well as climate change information that is easier to understand. To prevent media from being overwhelming and exacerbating climate anxiety, clear and concise information about climate change and the science behind it should be promoted in ways that contain limited jargon and are easy to understand for broader populations.

The results supported previous findings about eco-anxiety as a motivating factor for taking action through environmentally conscious behaviors and support of climate charities, but this effect is counterbalanced by the potential for worsened quality of life through damaged mental health and prevalence of discouraging emotions. The research built on previous findings by identifying specific negative impacts of ecoanxiety, such as trouble focusing and enjoying free time. The authors found that people who were anxious about climate change were more likely to hear about it frequently in the media, but felt information about climate change is overwhelming and hard to find and understand.

Limitations of the study include that the research was conducted only in English and only among US residents. Additionally, as the survey was conducted online, individuals without Internet access were unable to respond. The self-reported environmental and charitable behavior results may have been affected by events such as the COVID-19 pandemic, which prevented many people from volunteering in-person and potentially increased eco-anxiety rates. Furthermore, survey respondents had previously agreed to participate in research projects by being part of the survey panel used to conduct the reserach. These people may not fully represent the US population as a whole. Finally, the response rate, although high for an online survey, was not 100%. It is possible that the people who did not respond to the survey invitation are somehow different than those who did complete the survey.

The potential eco-anxiety solutions are based on self-reports about the factors that survey respondents thought would decrease their eco-anxiety; however, laypeople are often not the best predictors of their own future reactions (e.g., Poon, Koehler and Buehler, 2014). Even if actions that respondents felt could mitigate eco-anxiety would not immediately do so, they could nonetheless have societal benefits. Additionally, participation in social activities like volunteering could increase social cohesion and community connections, which previous research has shown have positive effects on psychological resilience and anxiety reduction, as well as other beneficial health outcomes (Bowe et al., 2021; Generaal et al., 2019; Howarth et al., 2020; Robinette et al., 2018; Robinette et al., 2021). Furthermore, the study did not cover personal experiences with climate-influenced weather changes. Demski et al. (2017) found that personal experience with extreme weather events like floods significantly increases people’s judgments of the importance of climate change as a social issue and their level of personal risk. Future research should further explore the relationship of perceived personal experience with climate-change-driven events, such as droughts or wildfires, with eco-anxiety.

## CONCLUSIONS

The results suggest that media exposure to climate change information was associated with higher likelihood of climate anxiety, but that self-defined level of knowledge about climate change was not significantly related to climate anxiety likelihood. These results may be related to eco-anxious individuals’ significantly higher likelihood to see climate change information as confusing, overwhelming, or hard to understand. Those with eco-anxiety were significantly more likely to indicate that more information about climate change’s causes, effects, and current efforts would reduce their anxiety. Developing and distributing straightforward, easy-to-understand climate change information may reduce eco-anxiety associated with media exposure while ensuring that these individuals remain accurately informed about the issue. More research is needed to explore the relationship between media exposure, knowledge, and eco-anxiety, as well as which information adaptations may most effectively reduce eco-anxiety and its association with media.

The findings support previous research about the positive and negative effects of eco-anxiety, and confirmed the hypothesis that anxiety about climate change could be reduced through behavioral strategies such as volunteering and information campaigns. It identified that eco-anxiety is a motivating factor encouraging people to take action, as well as several negative effects of eco-anxiety on the quality of life. The study expanded upon past research efforts that proposed behavioral strategies to reduce eco-anxiety by finding a significant association between volunteering efforts and predicted anxiety reduction. This is a novel solution, as volunteering for environmental organizations can reduce the negative effects of eco-anxiety while retaining motivation and interest and simultaneously have a positive impact on the environment. To encourage more people to volunteer, our study identified the importance of promoting information on how to get involved and encouraging newcomers. Future experimental research could test the impact of environmental volunteering to more clearly establish these effects.

The research also further explored the role of media and information on ecoanxiety, indicating that media about climate change can be overwhelming and contribute to higher rates of anxiety. To reduce this issue, it is important to cover climate change information in a way that is easy to understand by all populations. The media could also be used to promote information about how to get involved with local environmental volunteer groups, helping to encourage more people to get involved and reduce eco-anxiety. Media and online services can also be utilized to provide better climate mental health treatment access; Taylor (2020) notes that the COVID-19 pandemic has necessitated rapid online mental health service delivery, and that these advances could be adapted to help address climate anxiety.

Women and younger people were found to have a greater likelihood of ecoanxiety. Efforts and interventions to reduce eco-anxiety should be especially tailored towards women and young people to help combat this effect. The lack of a statistically significant relationship between education, political party membership, and eco-anxiety was unexpected and merits further study. Future research should also delve deeper into the association of eco-anxiety with age, gender, and other demographic factors. Potential changes to the experimental procedure would be to ask more questions surrounding age and youth perceptions of climate change, as previous studies identified that younger people were more likely to be eco-anxious, or questions targeting underrepresented racial, gender, and economic groups. Mixed-methods research could also be used to more deeply understand respondents’ experiences, attitudes and associations and possible reasons for them. Qualitative exploration has not yet been widely used in environmental epidemiology research, but has great promise for helping better understand causal processes and develop effective interventions (Noël, Vanroelen, & Gadeyne, 2021).

Furthermore, future studies should be conducted in multiple languages and with larger sample sizes to reduce the limitations of the study and further establish the robustness and reproducibility of the findings. Future research could also evaluate groups of people with and without eco-anxiety and assess the direct effects of different volunteering efforts or types of media exposure on anxiety reduction.

## Data Availability

All data produced in the present study are available upon reasonable request to the authors

## Ethical Standards

The authors assert that all procedures contributing to this work comply with the ethical standards of the relevant national and institutional committees on human experimentation and with the Helsinki Declaration of 1975, as revised in 2008. Institutional review was provided by WCG IRB. All survey respondents were adults aged 18+ who were informed of the objectives of the research and participated voluntarily. Formal consent was not obtained because survey data was collected anonymously.

## Availability of Data and Materials

Anonymized survey data is available upon request.

